# What delirium detection tools are used in routine clinical practice in the United Kingdom? Survey results from 91% of acute healthcare organisations

**DOI:** 10.1101/2021.01.12.21249699

**Authors:** Zoë Tieges, Jacqueline Lowrey, Alasdair M. J. MacLullich

## Abstract

**Purpose:** Our aim was to collect information on delirium assessment processes and pathways in non-intensive care settings in the United Kingdom (UK).

**Methods:** We sent a Freedom of Information request to 169 UK National Health Service (NHS) hospitals, trusts and health boards (units) in July 2020 to obtain data on usage of delirium assessment tools in clinical practice and delirium pathways or guidelines.

**Results:** We received responses from 154/169 units (91% response rate). Of these, 146/154 (95%) units reported use of formal delirium assessment processes and 131/154 (85%) units had guidelines or pathways in place. The 4 ‘A’s Test (4AT) was the most widely used tool, with 117/146 (80%) units reporting use. The Confusion Assessment Method was used in 65/146 (45%) units, and the Single Question to identify Delirium (SQiD) in 52/146 (36%) units.

**Conclusions:** Our findings show that the 4AT is the most commonly used tool in the UK, with 80% of units reporting use. This study adds to our knowledge of real-world uptake of delirium detection methods at scale. Future studies should evaluate real-world implementation of delirium assessment tools further via (i) tool completion rates and (ii) rates of positive scores against the expected prevalence delirium in the clinical population concerned.

**KEY SUMMARY POINTS:** *Aim:* To seek information on delirium assessment processes and pathways in non-intensive clinical care settings in the United Kingdom (UK), and to assess usage of specific delirium assessment tools: the 4 ‘A’s Test (4AT), Confusion Assessment Method and Single Question to identify Delirium (SQiD).

*Findings:* In total, 95% of National Health Service (NHS) units (hospitals, trusts and health boards) reported use of formal delirium assessment processes and 85% of units had guidelines or pathways in place. The 4AT was the most widely used tool, with 80% of units reporting use; the Confusion Assessment Method was reportedly used in 45% of units and the SQiD in 36% of units.

*Message:* This study shows real-world, large-scale uptake of delirium detection methods and delirium guidelines in UK hospitals, which contributes to ongoing efforts to improve delirium care.

## INTRODUCTION

Delirium is a serious acute neuropsychiatric disorder of arousal, attention and cognition [1]. It is independently associated with multiple poor outcomes, including higher mortality, new dementia, and patient and carer distress [1-4]. Delirium affects >15% of hospitalised patients, yet it remains under-detected in routine clinical practice [1, 5].

Detection is essential for the treatment of delirium, prompting the search for acute precipitants, and assessment and treatment of distress, managing delirium associated risks, and in communicating the diagnosis to patients and carers [1]. Formal detection of delirium in routine clinical practice at the earliest possible time point has been advocated in multiple guidelines. Considering the United Kingdom (UK), the 2010 National Institute of Clinical and Healthcare Excellence (NICE) guideline on delirium [6] recommended the Confusion Assessment Method (CAM), published in 1990 [7], for delirium assessment. NICE guidelines apply to England, Wales and Northern Ireland. Scotland’s main clinical guidelines are provided by the Scottish Intercollegiate Guidelines Network (SIGN); the 2019 SIGN delirium guidelines [8] recommended the 4 ‘A’s Test (4AT), a tool with the first validation study published in 2014 [9]. There is also a number of policies and standards promoting delirium detection, for example in hip fracture patients [10, 11]. Yet there is little evidence on what tools are in clinical use in the UK [1]. Knowledge of real-world practices on the use of such tools is essential in understanding of their implementability and may inform the content of future guidelines.

The aim of this study was to collect information on delirium assessment processes in clinical care in the UK, excluding Intensive Care Unit (ICU) settings, and to assess usage of specific delirium assessment tools for delirium detection. Specifically, we sought information on usage of two assessment tools recommended in clinical guidelines, the 4AT [12] and the CAM [7]. Use of the Single Question to identify Delirium (SQiD), a simple question used in some settings to screen for delirium by asking a friend or family member ‘Is this patient more confused than before?’, was also assessed. The SQiD was included because it has potential utility in non-specialist settings, either on its own or in combination with another assessment such as the 4AT if the SQiD is positive [13].

## METHODS

### Study design

Data on delirium assessment processes in hospitals across the UK were obtained through a Freedom of Information (FOI) request. The Freedom of Information Act 2000 provides access to information held by public authorities, including the National Health Service (NHS) [14].

The study authors constructed a short questionnaire composed of four questions (see Supplementary File 1) addressing usage of delirium assessment tools in clinical practice, specific tools used, clinical settings, and use of pathways or guidelines relating to delirium. Briefly, the questions addressed: (1) use of delirium assessment tool(s) as part of clinical practice for non-ICU patients; (2) which, if any, tools are used; (3) which, if any, validated tools are included in written policies; and (4) existence of pathways or guidelines relating to delirium. Trusts and health boards were asked to only include information collected before 31 July 2020.

The Freedom of Information Act was used to request data and no person identifiable data were sought. There were no aspects to this study requiring ethics committee approval.

The study was made available as a preprint (doi: https://doi.org/10.1101/2021.01.12.21249699).

### Data collection and analysis

NHS trusts and health boards (hereinafter termed ‘units’) are organisational units that may comprise one or more groups of hospitals. NHS units in the four UK countries England, Scotland, Wales and Northern Ireland were identified through lists available online. The FOI request was sent to generic email addresses (FOI and/or Information Governance teams) of 233 UK NHS units on 24 July 2020, with a reminder sent on 8 Oct 2020. Of these, 169 were either units with acute hospitals or non-acute hospital with geriatric rehab beds; here we report only on these units. Five were non-acute units providing inpatient rehabilitation care for older people; the other 164 units comprised acute hospitals or groups of hospitals. 143 units were based in England, 15 in Scotland, 6 in Wales and 5 in Northern Ireland. The majority were NHS trusts (England: N=142, Northern Ireland: N=5), 21 were regional NHS health boards (Scotland: N=15, Wales: N=6) and one was a hospital. The last date for responses was 30 Nov 2020. Descriptive analyses (number of responses, percentages) were performed using R Version 3.6.1 [15].

Data were collected by FOI respondents which are administrative staff who report on data held by their NHS unit and may consult with clinicians if they need to; it is beyond the scope of this study to take into account the differing levels of seniority of respondents.

## RESULTS

Responses to the FOI request were received from 154 out of 169 units (91% overall response rate) by 30 Nov 2020. This included responses from 149/164 acute units (91%) and from 5/5 (100%) non-acute units.

### Use of a delirium assessment tool as part of a formal delirium screening process in clinical practice

*(Questions 1 and 2)*

We asked units whether they used an assessment tool for delirium detection as part of clinical practice outside the ICU. In total, 146 out of 154 units (95%) reported use of a delirium assessment tool.

We also asked in which clinical settings these assessment tools were used. Delirium tool use in acute general medicine and/or geriatric medicine settings was reported by 126/146 (86%) units, in the Emergency Department by 94/146 (64%) units, and in surgical wards by 107/146 (73%) units (whereby some units specified the setting(s) in which a tool was used e.g. one unit specifically stated tool use in Orthopaedics).

### Validated delirium assessment tools included in written policies: 4AT, CAM, SQiD and/or other tools

*(Question 3)*

We asked units to state which of the following methods for delirium assessment were included in written (paper or electronic) policies: the 4AT, CAM, SQiD or another assessment tool. Of the 146 units which stated use of an assessment tool, 117/146 (80%) used the 4AT, 65/146 (45%) used the CAM, 52/146 (36%) used the SQiD and 7/146 (5%) used another tool.

Other tools or methods reported (one unit each) were: PINCHME, a mnemonic for the review of possible causes for delirium; the Observational Scale of Level of Arousal [16] for use in people with dementia; the National Early Warning Score (NEWS [17]); Recognizing Acute Delirium As part of your Routine (RADAR [18]); CAM for the Intensive Care Unit [19]; and one delirium assessment tool not further specified.

Of the 117 units reporting 4AT use, 77/117 (66%) stated that the 4AT was the only tool used, and 40/117 (34%) stated use of the 4AT alongside other tools. 14/117 (12%) units reported using 4AT as well as CAM (though not necessarily in the same setting) but not the SQiD or other assessment tool. 20/117 (17%) units reported using both the 4AT and the SQiD, but not the CAM or other tool. 20/117 (17%) units reported using the 4AT, the CAM and the SQiD.

Of the 65 units reporting CAM use, 26/65 (40%) stated that the CAM was the only tool used, whereas 39/65 (60%) stated use of CAM alongside other tools. 3/65 units (5%) reported using both the CAM and SQiD without other tools. Three units stated that the SQiD was the only delirium detection method used.

### Pathway or guidelines relating to delirium

#### (Question 4)

We asked units if they had a pathway or guidelines relating to delirium. 131/154 (85%) units stated that they had a delirium pathway or guidance in place. A further 11 acute units in England reported that such pathways or guidelines were under development.

## DISCUSSION

This UK-wide study with a high response rate of 91% found that 95% of units reported uptake of formal delirium assessment processes. In total, 85% of units had delirium guidelines or pathways in place. With respect to the tools used, the 4AT was the most widely used tool, with 80% reporting use, followed by the CAM in 45% of units and the SQiD in 36% of units. Several units reported using two or more of these tools; where only one tool was used, the 4AT was the most common, at 66%. The SQiD was generally not used alone, with only 3 units reporting use without another tool.

This study provides novel information on the national clinical uptake of delirium assessment tools. The UK has two main bodies producing clinical guidelines, NICE, covering England, Wales and Northern Ireland, and SIGN, covering Scotland. These bodies recommend the CAM and the 4AT, respectively, though importantly the 4AT was not published at the time that the NICE guidelines were published (2010 [6]). NICE recently stated that it will review its recommendations regarding delirium detection tools [20] in response to a randomised controlled trial showing that the 4AT had better sensitivity than the CAM with similar specificity [12]. This study and other factors such as the lack of need for special training for the 4AT (the CAM requires training), the relative brevity and clinical implementability of the 4AT, and the built-in cognitive testing in the 4AT may explain the greater uptake of the 4AT.

The SQiD is not directly comparable to the 4AT or CAM in that it is a brief single question used as an initial screen in advance of a more definitive tool such as the 4AT and CAM. It is of interest that around one third of UK hospitals report using the SQiD, despite the small evidence base supporting its use in hospital settings [21, 22]. The simplicity and high face validity of the SQiD may have led to its increasing adoption in the UK. Notably in April 2020 the Royal College of Physicians in the UK recommended using the SQiD in combination with the National Early Warning Score-2 as a method for monitoring new-onset delirium in hospital inpatients [23]. The guidance document states that if the SQiD is positive, the 4AT should then be done as a more definitive test.

This study has several strengths. Through use of the Freedom of Information process the response rate was high. This means that the findings are likely to provide a valid picture of delirium tool choice in UK hospitals. The findings thus reflect what individual units have chosen to use in clinical practice and the findings indicate some differences from the recommendations by UK guidelines committees. The study also revealed that 85% of UK units have formal delirium pathways in place; this was not previously known. Some limitations should be acknowledged. The study did not provide information on usage rates of the tools, or diagnostic performance of the tools in real-world practice; this is important to note because reporting that delirium detection tools *should* be used does not necessarily mean that these tools *are* used in routine practice. Information on characteristics of units was not sought, which precluded opportunities for further stratified analyses; however, the questionnaire was deliberately kept short to maximise response rate. Though we sourced all known contact emails for UK units it is possible that some units were missed. However, the study is likely to have reported on the vast majority of units.

Future work should examine uptake of delirium detection tools in different settings and other countries. Alongside this, information on two key implementation parameters should be collected: (a) completion rates in eligible populations, and (b) rates of positive scores in completed tests assessed against the expected rates of delirium in the clinical population concerned [5]. This pair of metrics is essential to understand real-world value and use of delirium tools, and gathering such data is increasingly feasible through large scale electronic health records. As one example, the 4AT is mandated for all hip fracture patients in England and whole clinical population data from 2017 (total n = 60,000 patients) showed that 95% of patients were screened with the 4AT, with 25% having a positive score [11].

Our findings showed that 12% of NHS units reported using the 4AT as well as the CAM, though it was not ascertained if these tools were used in the same or different clinical settings (or patients). It would be of interest in future studies to better understand potential advantages and pitfalls of using more than one delirium assessment tool in the same setting, or even in the same patient at different times.

In conclusion, this study provides novel information on the reported uptake of delirium detection tools in >90% of UK hospitals. The 4AT is the most commonly used tool, with 80% of units reporting implementation. This study adds to our knowledge of real-world large-scale implementation of delirium detection methods, and contributes to ongoing efforts to improve delirium care and to develop awareness of the importance of delirium screening within the UK and globally.

**Table 1.**
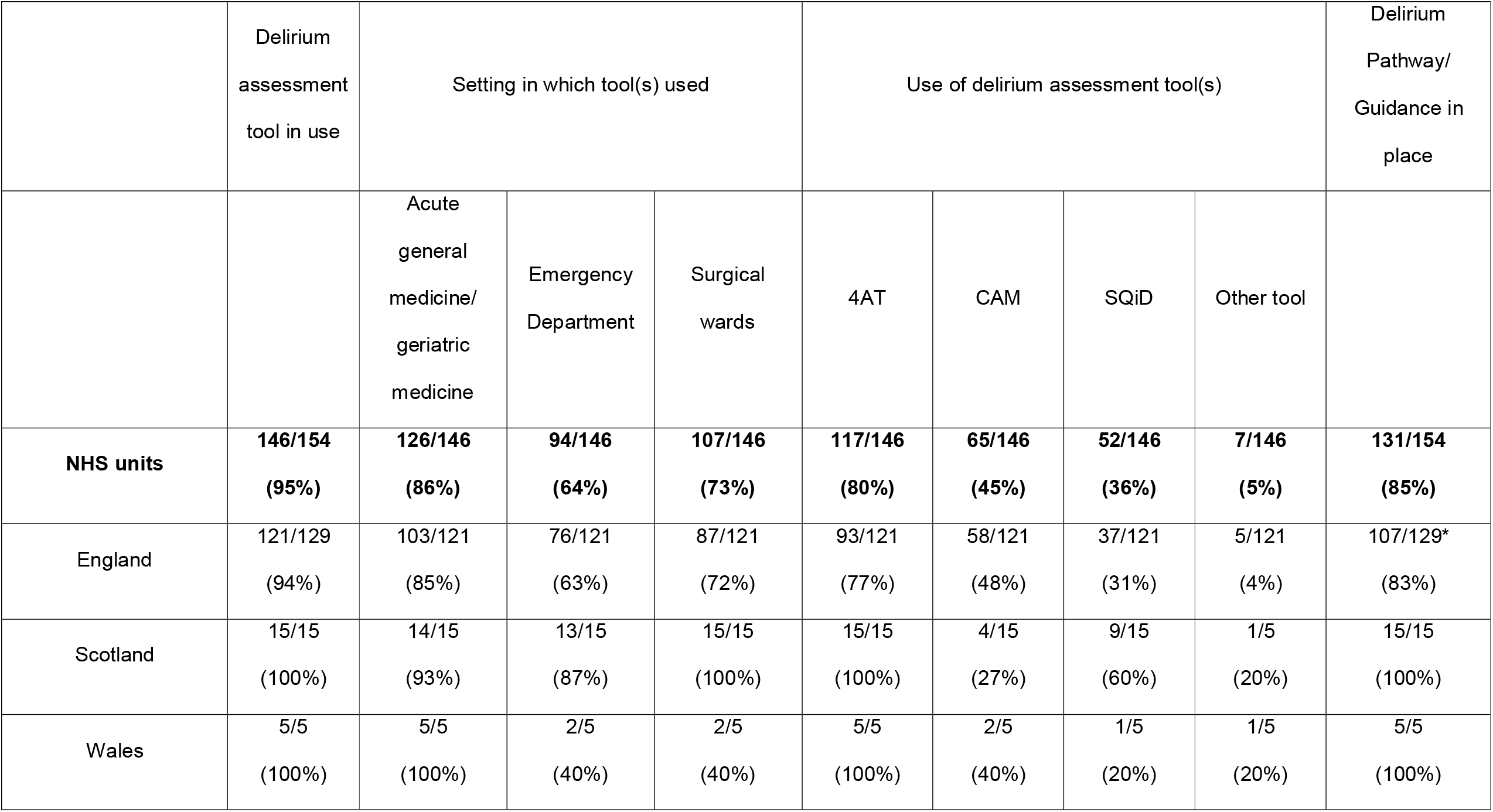
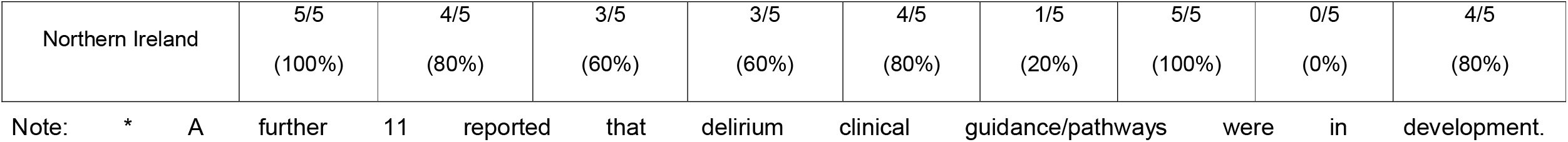
Results from Freedom of Information request on delirium assessment in UK hospitals. NHS: National Health Service. CAM: Confusion Assessment Method. SQiD: Single Question in Delirium.

|  | Delirium<br>assessment<br>tool in use | Setting in which tool(s) used |  |  | Use of delirium assessment tool(s) |  |  |  | Delirium<br>Pathway/<br>Guidance in<br>place |
| --- | --- | --- | --- | --- | --- | --- | --- | --- | --- |
|  |  | Acute<br>general<br>medicine/<br>geriatric<br>medicine | Emergency<br>Department | Surgical<br>wards | 4AT | CAM | SQID | Other tool |  |
| **NHS units** | **146/154**<br>**(95%)** | **126/146**<br>**(86%)** | **94/146**<br>**(64%)** | **107/146**<br>**(73%)** | **117/146**<br>**(80%)** | **65/146**<br>**(45%)** | **52/146**<br>**(36%)** | **7/146**<br>**(5%)** | **131/154**<br>**(85%)** |
| England | 121/129<br>(94%) | 103/121<br>(85%) | 76/121<br>(63%) | 87/121<br>(72%) | 93/121<br>(77%) | 58/121<br>(48%) | 37/121<br>(31%) | 5/121<br>(4%) | 107/129*<br>(83%) |
| Scotland | 15/15<br>(100%) | 14/15<br>(93%) | 13/15<br>(87%) | 15/15<br>(100%) | 15/15<br>(100%) | 4/15<br>(27%) | 9/15<br>(60%) | 1/5<br>(20%) | 15/15<br>(100%) |
| Wales | 5/5<br>(100%) | 5/5<br>(100%) | 2/5<br>(40%) | 2/5<br>(40%) | 5/5<br>(100%) | 2/5<br>(40%) | 1/5<br>(20%) | 1/5<br>(20%) | 5/5<br>(100%) |
| Northern Ireland | 5/5<br>(100%) | 4/5<br>(80%) | 3/5<br>(60%) | 3/5<br>(60%) | 4/5<br>(80%) | 1/5<br>(20%) | 5/5<br>(100%) | 0/5<br>(0%) | 4/5<br>(80%) |

Note: \* A further 11 reported that delirium clinical guidance/pathways were in development.

## Data Availability

Information gained through FOI Act is publicly available.

## Acknowledgments

The authors would like to thank Sharon Moncrieff and Maureen Harding who were responsible for the distribution of the Freedom of Information requests and data collection.

## DECLARATIONS

### Availability of data and material

Information gained through the FOI Act is publicly available.

### Code availability

Not applicable.

### Authors’ contributions

All authors were involved in the conception and design of the Freedom of Information request and study. ZT conducted data analysis with input from AMJM and JL. ZT and AMJM drafted the manuscript. All authors critically appraised the manuscript, and read and approved the final manuscript. AMJM and JL were involved in acquiring the funding for this research.

## Supplementary File 1: Freedom of Information questions

1. Do you use a delirium assessment tool as part of clinical practice for your non-ICU patients in your trust/hospital? YES / NO
2. If yes, in which clinical settings are they in place (please use X to indicate all that apply)? _________ Acute general medicine/Medicine of the Elderly _________ Emergency Department _________ Surgical wards _________ Other (please specify):
3. Which, if any, validated tools are included in your written (paper or electronic) policies? Please use X to indicate all that apply. _________ 4 ‘A’s Test (4AT) _________ Confusion Assessment Method _________ Single Question in Delirium _________ Other (please specify):
4. Do you have a pathway or guidelines relating to delirium? YES / NO If yes, in which year were they written? Please attach an electronic copy.

